# Intraoperative effects of ETV and CPC on intraventricular pressure and pulsation amplitude: A preliminary investigation of the hydrodynamic model of infant hydrocephalus

**DOI:** 10.64898/2026.06.24.26355729

**Authors:** Marcia H. Yoshikawa, German Figueroa, M. Elena Dominguez-Villasenor, P. Ellen Grant, Jason Sutin, Benjamin C. Warf, Pei-Yi Lin

**Affiliations:** Fetal-Neonatal Neuroimaging and Developmental Science Center, Boston Children’s Hospital, 300 Longwood Ave., Boston, MA 02115, USA; Division of Newborn Medicine, Department of Pediatrics, Boston Children’s Hospital, 300 Longwood Ave., Boston, MA 02115, USA; Harvard Medical School, 25 Shattuck St., Boston, MA 02115, USA; Department of Radiology, Boston Children’s Hospital, 300 Longwood Ave., Boston, MA 02115, USA; Department of Neurosurgery, Boston Children’s Hospital, 300 Longwood Ave., Boston, MA 02115, USA

**Keywords:** Hydrocephalus, Infant hydrocephalus, Endoscopic third ventriculostomy, Choroid plexus cauterization, Intracranial pressure, Hydrodynamic model, Intraventricular pulsation

## Abstract

**Background:** The hydrodynamic model of hydrocephalus proposes that ventriculomegaly is driven by exaggerated intraventricular pulsations rather than impaired cerebrospinal fluid (CSF) circulation alone. Under this model, endoscopic third ventriculostomy with choroid plexus cauterization (ETV/CPC) treats hydrocephalus by creating a pulsation absorber and by reducing a primary source of intraventricular pulsation. However, direct intraoperative human evidence supporting this two-step mechanism is lacking. This study aimed to test the hypothesis that ETV followed by CPC would produce measurable, stepwise decreases in mean intraventricular pressure (ICP) and pulsation amplitude in infants with hydrocephalus.

**Methods:** This single-institution proof-of-concept study included infants with symptomatic hydrocephalus undergoing ETV/CPC as the first definitive treatment. A fiber-optic ICP sensor was attached to the operative ventriculoscope and passively recorded mean and pulsatile ICP (pulsation amplitude) throughout the procedure. Longitudinal brain parenchymal volume (BPV) and cerebrospinal fluid volume (CSFV) were obtained through segmentation of clinically acquired T2-weighted MRI and converted to age– and sex-matched z-scores. All patients were followed for a minimum of 6 months postoperatively.

**Results:** Five infants (median corrected age at ETV/CPC 8 months) were included. No surgical complications occurred, and no ETV/CPC failures were observed during follow-up. Overall, mean ICP decreased by 56–97% after the combined procedure in four patients. In three patients (Patients 1, 3, and 5), both mean ICP and pulsation amplitude decreased stepwise following ETV and then CPC, consistent with the hypothesized therapeutic mechanism. Patient 4 demonstrated a large reduction in mean ICP after ETV with minimal additional effect from CPC and no significant change in pulsation amplitude. Patient 2 demonstrated neither a reduction in mean ICP nor a meaningful change in pulsation amplitude after either procedure; this patient also had a delayed and atypical clinical response. Intracranial segmentation demonstrated BPV z-score stabilization within normal range and CSFV plateau in all patients after surgery.

**Conclusions:** This proof-of-concept study provides the first direct intraoperative human evidence supporting the hydrodynamic mechanism of ETV/CPC in a subset of infant with hydrocephalus. Our findings suggest that determination of intraoperative ICP parameters is feasible, safe and might ultimately prove helpful in improving patient selection for ETV/CPC, warranting further investigation in larger cohorts.

## INTRODUCTION

Since the early 1960s, shunting the cerebrospinal fluid (CSF) from cerebral ventricles to other body cavities has been the standard treatment for hydrocephalus. However, shunts are prone to repeated failures, with nearly half failing at least once within two years and the majority failing over the longer term [1–4]. Progress in developing alternative treatments has been slow due to an incomplete understanding of CSF physiology and the complex processes that drive hydrocephalus [5–6].

Traditionally, hydrocephalus has been explained as a disorder of CSF circulation in which interruption of flow between the choroid plexus and the arachnoid granulations leads to ventricular enlargement, known as the bulk flow model [7]. However, several observations challenge the adequacy of this model, especially in infants: arachnoid granulations play a minimal role in CSF absorption at this age, intraventricular CSF flow is bidirectional, and CSF egress through the periventricular white matter into the glymphatic system has been demonstrated [8, 9]. Taken together, these findings indicate that bulk flow alone is insufficient to account for the pathophysiology of infant hydrocephalus, motivating the search for an alternative framework.

The hydrodynamic model is an alternative explanation for hydrocephalus development by proposing that it is a disorder of the intracranial pulsations rather than a disturbance of CSF flow and absorption. In this model, arterial systolic pressure waves delivered to the intracranial compartment by the vertebral, basilar, and carotid arteries are initially dampened by the Windkessel mechanism of the subarachnoid space before entering the brain parenchyma. Intracranial pulsation absorption is accomplished by the compliant cerebral venous system. The highly vascularized choroid plexus delivers systolic pulsations into the intraventricular CSF which are dampened by means of the pulsatile outflow of CSF from the ventricles into the subarachnoid spaces. Experimentally, progressive ventriculomegaly has been produced by augmenting the intraventricular pulsation amplitude despite normal mean ICP [10, 11]. Conversely, intraventricular pulsations were dampened and progressive ventriculomegaly prevented in an animal model of hydrocephalus in the face of elevated intraventricular pressure by the removal of choroid plexus [10, 11]. Thus, exaggerated intraventricular pulsations appear to help drive progressive ventriculomegaly. Congenitally or secondarily disabled mechanisms of pulsation absorption (e.g. dysfunctional subarachnoid space, decreased venous compliance, or ventricular outlet obstruction) may increase the intraventricular pulsation amplitude and lead to progressive ventriculomegaly. [5, 6, 10–13]

Endoscopic third ventriculostomy (ETV) is a treatment for hydrocephalus that can avoid life-long dependence upon ventricular shunts for many patients. But ETV is less successful for infants than it is for older children. The rate of success for infants is much improved with the addition of choroid plexus cauterization (CPC) in which the choroid plexus in both lateral ventricles is effectively removed [4, 5, 14–20]. An interpretation of the hydrodynamic model of hydrocephalus suggests that ETV serves as a pulsation absorber, reducing the intraventricular pulsation amplitude. One possible explanation as to why this is less effective for infants is based upon the observation that infants have a higher brain compliance due to incomplete myelination [12] suggesting that the ventricles expand more readily, rendering the pulsation absorber effect of ETV insufficient in some cases. This explanation is supported by our observation that two independent factors influence the likelihood of ETV/CPC success: age and ventricle size [4, 21, 22]. The rate of success is lower with decreasing age and with increasing ventricular volume [4, 21, 22], both of which correlate with increased brain compliance. Our hypothesis has been that the addition of CPC may further diminish the intraventricular pulsation amplitude, as demonstrated in previous animal models [13, 23], thus augmenting the pulsation absorber effect of the ETV sufficiently to increase the rate of success.

Despite decades of clinical experience with ETV/CPC, its proposed therapeutic mechanism has never been directly tested in humans intraoperatively. Prior supporting evidence derives entirely from animal models, computational simulations, and indirect clinical observations. Here, we describe a novel, minimally invasive method to co-register ICP continuously during the operative procedure using a fiber-optic sensor integrated into the ventriculoscope without modification of or interference with the surgical technique. This approach enabled, for the first time, direct measurement of the stepwise effects of ETV and CPC on mean and pulsatile ICP in human infants.

## METHODS

This pilot investigation was performed at a single institution and received ethical approval from the institutional review board at Boston Children’s Hospital. The guardians of all included patients provided written consent to participate in this study.

### Methodological Innovation

A key feature of this study was the integration of continuous intraoperative ICP monitoring into the standard ETV/CPC procedure without any modification to the operative technique. The fiber-optic ICP sensor was assembled passively into the existing ventriculoscope via a Tuohy Borst adapter alongside the standard cauterization electrode and irrigation tubing, adding no additional operative steps, instruments, or procedural risk to the patient. This approach preserves the standard surgical workflow in its entirety while enabling real-time pressure acquisition throughout the procedure, a critical prerequisite for clinical adoption at scale.

### Patient Selection

Patients with symptomatic hydrocephalus, corrected age less than 24 months, and undergoing ETV/CPC as the first definitive treatment for hydrocephalus were eligible for this study.

### Intraoperative ICP Monitoring Procedure

In the sterile field, a single-use fiber-optic ICP sensor (FISO Technologies, Québec, Canada) was attached to the 3.7-mm flexible steerable ventriculoscope (Karl Storz, Tuttlingen, Germany) as detailed in Figures 1 and 2. The ICP sensor, the clinical cauterization electrode, and a semirigid tube attached to a syringe were inserted into the ventriculoscope through the Tuohy Borst adapter, which allowed adjustment of the sensor and electrocauterization wire during surgery while maintaining a fluid seal for ICP monitoring. The ventriculoscope and assembly were flushed with saline prior to use. Once assembled, ICP was passively recorded throughout the entire procedure. It is important to note that no CSF escaped or was aspirated nor was irrigation used at any point throughout the procedure, as such would have contributed to any observed changes in pressure and/or pulsatility. At each timepoint of interest (baseline, post-ETV, and post-CPC), a one-minute pause was observed to allow ICP recording with minimal motion noise. The surgical team was blinded to ICP values throughout the procedure, ensuring that operative decisions were made independently of the monitoring data and that no performance bias was introduced.

**Figure 1.**
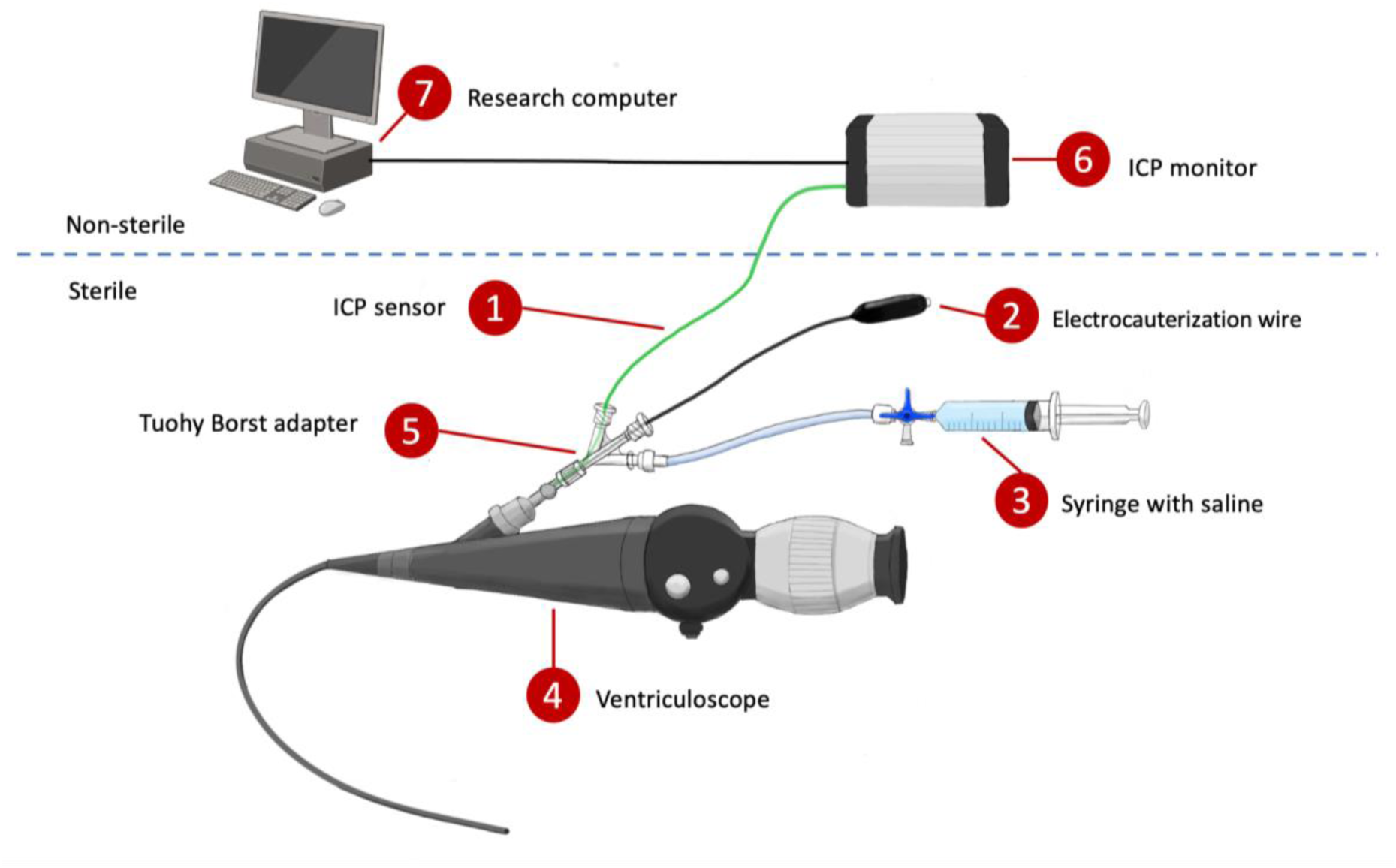
Schematic illustration of the intraoperative intracranial pressure (ICP) monitoring setup. The ICP sensor (1), the clinical electrocauterization wire (2), and a syringe filled with saline (3) are attached to the ventriculoscope (4) though a Tuohy Borst adapter (5). The ICP sensor is connected to an ICP monitor (6)(outside the sterile field), which, in its turn, is connected to a research computer (7).

**Figure 2.**
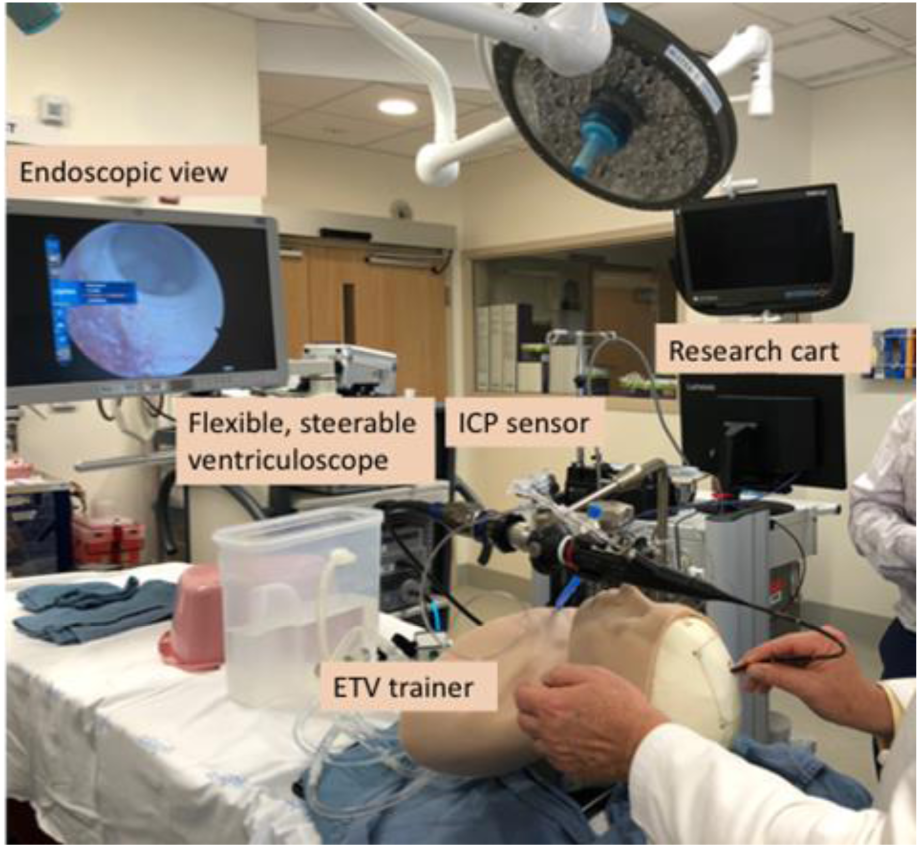
Simulation of intraoperative ICP measurement with the clinical and research devices in a custom ETV trainer in a simulated operating room at Boston Children’s Hospital’s SIMPeds. ICP: intracranial pressure; ETV: endoscopic third ventriculostomy.

### Data Collection and Analysis

The mean and pulsatile ICP were computed using a previously published method [24]. Clinical and surgical data were obtained from medical records. Clinically acquired axial T2-weighted MRI scans were retrieved and segmented to obtain brain parenchymal volume (BPV) and cerebrospinal fluid volume (CSFV). MRI protocols included HASTE, STEALTH, and PROPELLER. Intracranial segmentation was performed using a deep-learning model tailored for the infant hydrocephalus population [25]. Segmentation results were quality-checked and manually edited by trained staff (M.H.Y.) using ITK-SNAP [26] and verified by a board-certified pediatric neuroradiologist (P.E.G.).

To account for age– and sex-related changes in BPV and CSFV, z-scores were calculated based on normative growth curves from Peterson et al [27]. Because infants with hydrocephalus can have intracranial volumes far outside the range of healthy normative data, standard z-score methods produce numerically unstable or truncated results at these extremes; an updated computation was therefore applied to generate unbounded z-scores capable of representing such outlying values without distortion. This was implemented using the generalized method proposed by Rigby [28], computed in log-space to ensure numerical stability across the full range of observed values.

## RESULTS

Five infants were enrolled in this study (Table 1). All patients underwent successful ETV/CPC with greater than 90% cauterization of the choroid plexus in both lateral ventricles. No intraoperative or postoperative complications occurred, and no adverse events were attributable to the ICP monitoring setup. All patients were followed for at least 6 months postoperatively (range 6–12 months) and no ETV/CPC failures were identified during the follow-up period.

**Table 1.** Baseline information of included patients. ETV/CPC: endoscopic third ventriculostomy with choroid plexus cauterization; CSFV: cerebrospinal fluid volume; BPV: brain parenchymal volume.

|  | Patient #1 | Patient #2 | Patient #3 | Patient #4 | Patient #5 |
| --- | --- | --- | --- | --- | --- |
| **Sex** | Male | Male | Female | Female | Male |
| **Premature** | Yes | No | Yes | No | Yes |
| **Preop CSFV z-score** | 5.87 | 3.92 | 5.30 | 5.40 | 6.68 |
| **Preop BPV z-score** | 2.38 | 1.32 | 0.34 | 1.72 | -0.15 |
| **Additional MRI findings** | Enlarged posterior fossa; left middle cranial fossa arachnoid cyst | Enlarged posterior fossa; midline arachnoid cyst elevating the floor of the third ventricle | Chiari II; absent septum pellucidum | Enlarged posterior fossa | Chiari II |

### Patient Descriptions

**Patient 1.** A male infant with corrected age between 6 and 12 months presented with accelerated head circumference growth and downward gaze preference. Serial MRIs demonstrated progressive ventricular enlargement. Additional findings included a prominent retrocerebellar CSF space in communication with the fourth ventricle with widening of the tegmento-vermian angle, a left middle cranial fossa arachnoid cyst, patent cerebral aqueduct, and an intact cerebellar vermis. He underwent ETV/CPC without complications. At 12-month postoperative follow-up, ventricular size had decreased and head circumference growth had slowed. The patient showed no signs of elevated intracranial pressure and was meeting developmental milestones appropriately. Intracranial segmentation demonstrated BPV z-scores within the normal range and postoperative stabilization of CSFV z-scores (Figure 3A).

**Figure 3.**
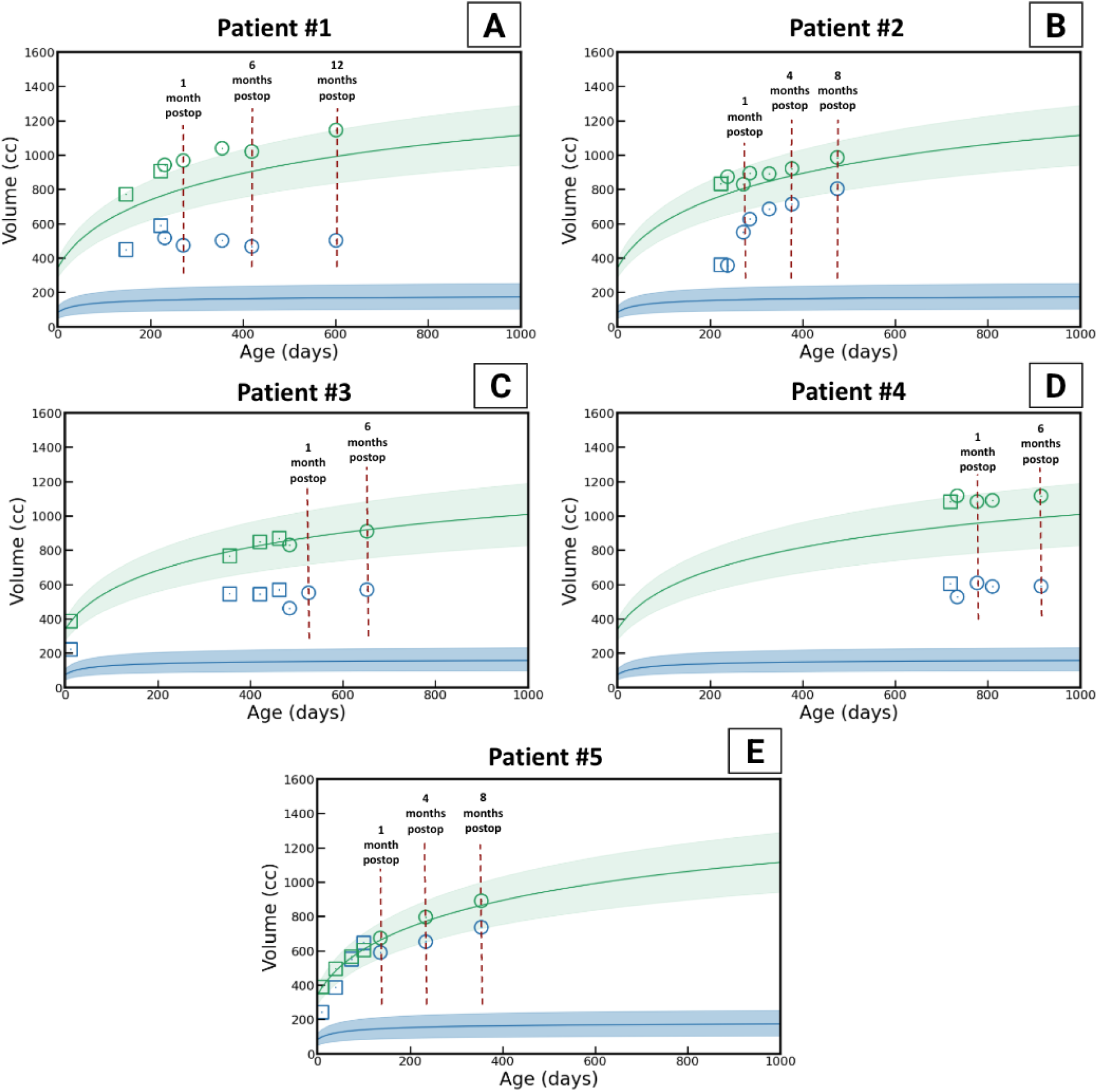
Trajectories of brain parenchyma and cerebrospinal fluid (CSF) volumes of patients 1 (A), 2 (B), 3 (C), 4 (D), and 5 (E). Brain (green curve) and CSF (blue curve) volumes are plotted as a function of age (days). Individual data points represent preoperative (squares) and postoperative (circles) measurements. Solid lines indicate normative growth curves, and shaded regions denote the ±2 standard deviation (z-score) normative range.

**Patient 2.** A male infant with corrected age between 6 and 12 months and no significant past medical history presented with accelerated head circumference growth and an enlarged fontanelle. Brain MRI demonstrated severe supratentorial and infratentorial ventriculomegaly with distension of the fourth ventricle outlet foramina and the cerebral aqueduct. A pre-pontine cistern arachnoid cyst was identified elevating the floor of the third ventricle and displacing the brainstem posteriorly and the pituitary stalk anteriorly; this finding was reviewed by the treating neurosurgeon (B.W.) and neuroradiologist (P.E.G.) and was not considered to interfere with the technical execution or expected efficacy of the ETV. The patient underwent cyst fenestration and ETV/CPC without procedural complication.

Postoperatively, ventricular volume as determined by frontal-occipital horn ratio (FOHR) continued to increase, as did head circumference z-score for the first three months. The patient remained clinically well throughout with a flat to sunken fontanelle and no symptoms of elevated intracranial pressure; however, given the progressive ventriculomegaly and persistent head growth, placement of a VP shunt was planned because of presumed treatment failure. However, at 4 months post-ETV/CPC, neither the ventricular size (as determined by FOHR) nor the head circumference z-score had increased since the 3-month evaluation. In view of this improvement and his good clinical condition, shunt placement was deferred and the patient has since continued to do well. Intracranial segmentation demonstrated BPV z-scores within the normal range; CSFV z-scores increased in the initial postoperative period before slowing its growth at approximately 4 months after surgery, consistent with the clinical trajectory described above (Figure 3B).

**Patient 3.** A female with corrected age between 13 and 18 months, Chiari II malformation and myelomeningocele presented with accelerated head circumference growth and delays in motor and speech development. Her past medical history was significant for in utero myelomeningocele repair and preterm delivery. MRI demonstrated ventriculomegaly of the lateral and third ventricles and absence of the septum pellucidum. She underwent ETV/CPC and had an uneventful recovery. At 6-month follow-up there were no signs of ETV/CPC failure. Intracranial segmentation demonstrated normal BPV z-scores and stabilization of CSFV z-scores (Figure 3C).

**Patient 4.** A female with corrected age between 19 and 24 months presented with macrocephaly and gross motor developmental delay. MRI revealed ventriculomegaly involving the lateral, third, and fourth ventricles as well as diffuse thinning of the white matter. No obstructing lesions were identified. She underwent ETV/CPC and tolerated the procedure well without complication. At 6-month follow-up, there were no signs of ETV/CPC failure. Intracranial segmentation demonstrated normal BPV z-scores and stable CSFV z-scores (Figure 3D).

**Patient 5.** A male infant with corrected age < 6 months, prenatally diagnosed with Chiari II malformation and myelomeningocele was identified on routine surveillance MRI to have progressive supratentorial ventriculomegaly. Notably, Patient 5 was the youngest in this study and had the highest preoperative CSFV z-score among all patients. He underwent ETV/CPC and had an uneventful recovery. At 8-month post-operative follow-up, there were no signs of ETV/CPC failure. Intracranial segmentation demonstrated normal BPV z-scores and stable CSFV z-scores (Figure 3E). Patient 5 demonstrated substantial amount of intracranial air within the subarachnoid spaces in postoperative imaging (Figure 5). Consequently, the apparent increase in CSFV observed during follow-up likely reflects reabsorption of postoperative air rather than expansion of the CSF-filled ventricular or subarachnoid spaces (Figure 3E).

### Intraoperative ICP Monitoring Results (Table 2)

Baseline mean ICP varied considerably across patients, ranging from 3.0 mmHg (Patient 3) to 24.7 mmHg (Patient 1), with Patient 1 having the highest baseline mean ICP in this study (Table 2, Figure 4A). The remaining four patients had baseline mean ICP values between 3.0 and 11.8 mmHg. Overall, four of five patients (Patients 1, 3, 4, and 5) demonstrated a substantial decrease in mean ICP following the combined ETV/CPC procedure (56–97% total reduction). Patient 2 was the outlier, with only a 4% total decrease in mean ICP across both procedural steps.

**Figure 4.**
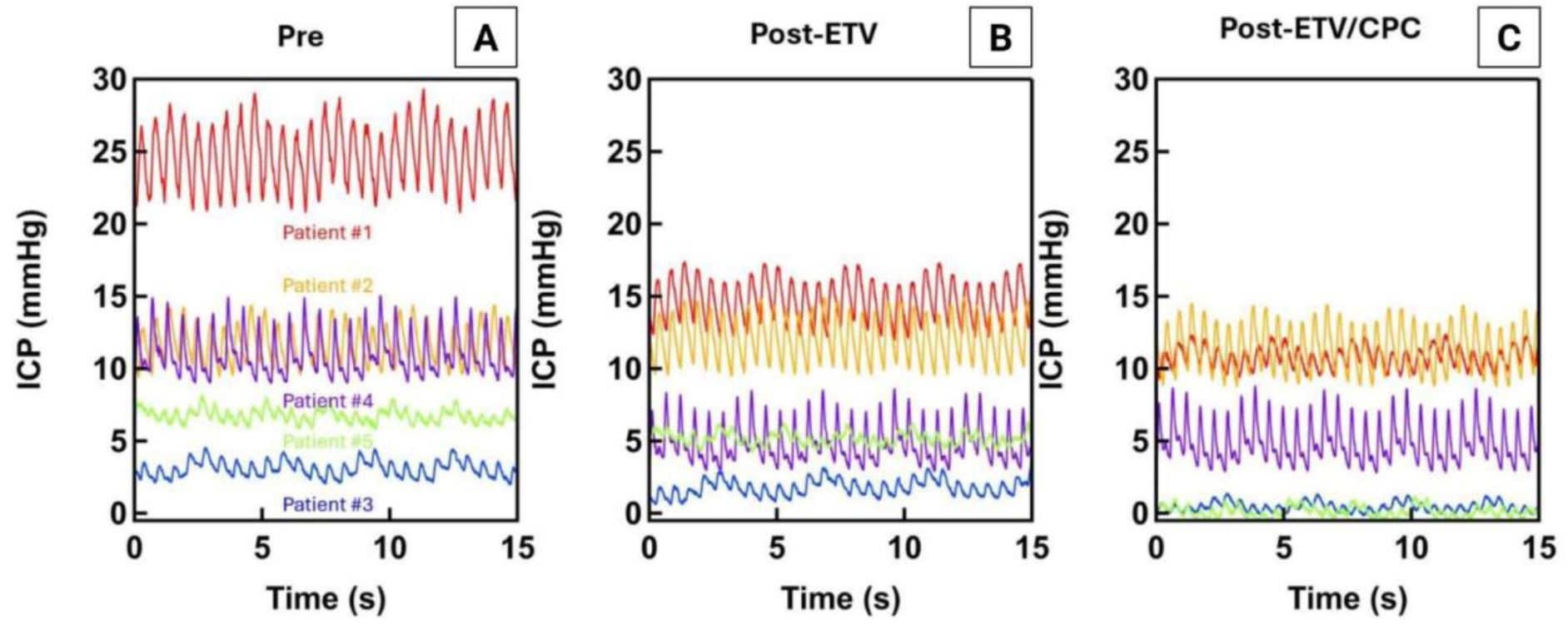
Mean ICP and pulse ICP change at baseline (A), post-ETV (B), and post-CPC (C). ETV: endoscopic third ventriculostomy; CPC: choroid plexus cauterization; ICP: intracranial pressure.

**Figure 5.**
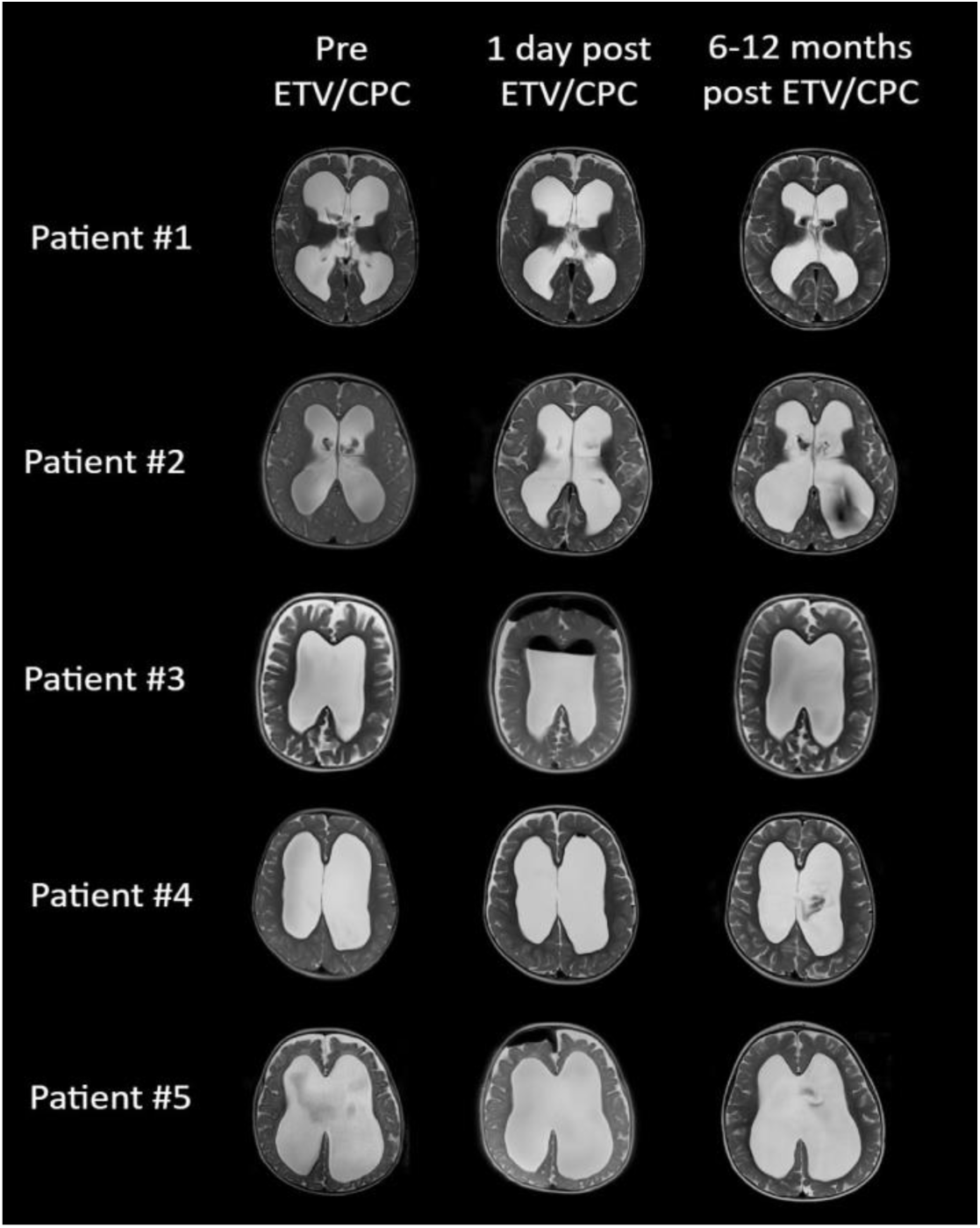
Axial T2 Haste MRI from patients at different timepoints. Each image represents the slice with the largest diameter of the lateral ventricles on the axial view. ETV/CPC: endoscopic third ventriculostomy with choroid plexus cauterization.

**Table 2.**
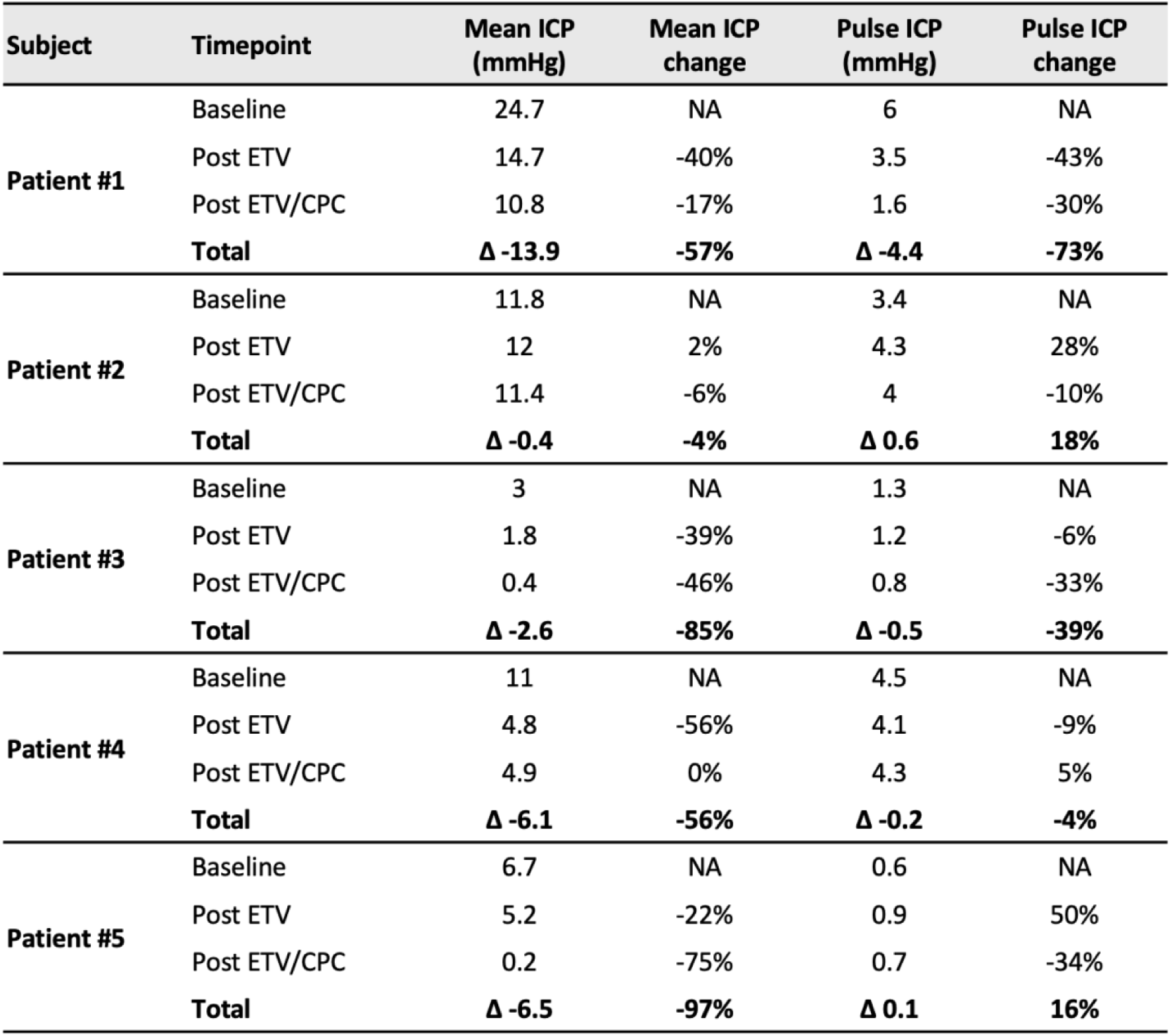
Mean and pulse intracranial pressure (ICP) during the surgical procedure. ETV: endoscopic third ventriculostomy; CPC: choroid plexus cauterization.

| Subject | Timepoint | Mean ICP (mmHg) | Mean ICP change | Pulse ICP (mmHg) | Pulse ICP change |
| --- | --- | --- | --- | --- | --- |
| **Patient #1** | Baseline | 24.7 | NA | 6 | NA |
|  | Post ETV | 14.7 | -40% | 3.5 | -43% |
|  | Post ETV/CPC | 10.8 | -17% | 1.6 | -30% |
|  | **Total** | **Δ -13.9** | **-57%** | **Δ -4.4** | **-73%** |
| **Patient #2** | Baseline | 11.8 | NA | 3.4 | NA |
|  | Post ETV | 12 | 2% | 4.3 | 28% |
|  | Post ETV/CPC | 11.4 | -6% | 4 | -10% |
|  | **Total** | **Δ -0.4** | **-4%** | **Δ 0.6** | **18%** |
| **Patient #3** | Baseline | 3 | NA | 1.3 | NA |
|  | Post ETV | 1.8 | -39% | 1.2 | -6% |
|  | Post ETV/CPC | 0.4 | -46% | 0.8 | -33% |
|  | **Total** | **Δ -2.6** | **-85%** | **Δ -0.5** | **-39%** |
| **Patient #4** | Baseline | 11 | NA | 4.5 | NA |
|  | Post ETV | 4.8 | -56% | 4.1 | -9% |
|  | Post ETV/CPC | 4.9 | 0% | 4.3 | 5% |
|  | **Total** | **Δ -6.1** | **-56%** | **Δ -0.2** | **-4%** |
| **Patient #5** | Baseline | 6.7 | NA | 0.6 | NA |
|  | Post ETV | 5.2 | -22% | 0.9 | 50% |
|  | Post ETV/CPC | 0.2 | -75% | 0.7 | -34% |
|  | **Total** | **Δ -6.5** | **-97%** | **Δ 0.1** | **16%** |

**Patients 1 and 3** demonstrated the response pattern following ETV and then CPC that is most consistent with the hydrodynamic model (ETV acting as a pulsation absorber and CPC functioning to further diminish the pulsation amplitude) with a significant stepwise decrease in both mean ICP and pulsation amplitude in response to each procedural component (Figure 4). Patient 1 showed a 40% decrease in mean ICP after ETV (24.7→14.7 mmHg) and a further 17% decrease after CPC (14.7→10.8 mmHg), with pulsation amplitude decreasing 43% after ETV and a further 30% after CPC. Patient 3 showed a 39% decrease in mean ICP after ETV (3.0→1.8 mmHg) and a further 46% decrease after CPC (1.8→0.4 mmHg), with pulsation amplitude decreasing 6% after ETV and a further 33% after CPC. Notably, the mean ICP for Patient 3 following ETV/CPC approached zero (0.4 mmHg), whereas Patient 1 retained a post-ETV/CPC mean ICP of 10.8 mmHg despite a substantial overall reduction.

**Patient 5** demonstrated a 22% decrease in mean ICP following ETV (6.7→5.2 mmHg), with a further substantial decrease of 75% after CPC (5.2→0.2 mmHg), resulting in a post-ETV/CPC mean ICP approaching zero, paralleling the finding in Patient 3. As noted, this patient was the youngest in this study and had the highest preoperative CSFV z-score of 6.68. Baseline pulsation amplitude in Patient 5 was the lowest of all patients (0.6 mmHg). Although a relative increase of 50% in pulsation amplitude was observed after ETV (0.6→0.9 mmHg), with a subsequent decrease of 34% after CPC (0.9→0.7 mmHg), the absolute values throughout were negligible.

**Patient 4** exhibited the largest proportional decrease in mean ICP after ETV alone in this study (56%; 11.0→4.8 mmHg). However, no further decrease in mean ICP occurred after CPC (4.8→4.9 mmHg, 0% change). Pulsation amplitude showed minimal change at both steps (−9% after ETV; +5% after CPC), with a net minor decrease of 4% on pulsatility.

**Patient 2** was the notable exception in this study. Unlike the other 4 patients, mean ICP showed no significant change following ETV (+2%; 11.8→12.0 mmHg) and only a minor decrease after CPC (−6%; 12.0→11.4 mmHg), for a total reduction of 4%. Surprisingly, the pulsation amplitude increased by 28% after ETV (3.4→4.3 mmHg), followed by a partial recovery of −10% after CPC (4.3→4.0 mmHg), resulting in a net increase of 18% above baseline at the end of the procedure. This patient was also unique in that there was an arachnoid cyst in the prepontine cistern that was simultaneously fenestrated at the time of the ETV. It is not at all apparent, however, that this would have accounted for the aberrant responses in ICP and pulsatility. It is of interest that this was the only patient who had a delayed response to treatment.

### Trajectories of Brain Parenchyma and Cerebrospinal Fluid Volumes

Intracranial segmentation analysis (Figure 3) demonstrated stabilization of BPV z-scores within the normal range following surgery in all patients. CSFV z-scores plateaued in four of five patients following ETV/CPC, although remaining above normative values for age and sex in healthy children. Patient 2 demonstrated an initial increase in CSFV z-scores in the early postoperative period, followed by a plateau at approximately 4 months after surgery. Axial T2-weighted HASTE MRI images obtained at 2 weeks prior to surgery, one day after ETV/CPC, and at last postoperative follow-up are shown in Figure 5. The timing of the last MRI ranged from 6 to 12 months after surgery: 12 months for Patient 1, 8 months for Patient 2, and 6 months for Patients 3, 4, and 5.

### Relationship between ICP and CSF Volume

Inspection of Figure 6 reveals that neither baseline mean ICP nor baseline pulsation amplitude demonstrated a clear or consistent correlation with preoperative CSFV across patients (Figure 5C–D). The patient with the largest preoperative CSFV z-score (Patient 5) had the lowest baseline pulsation amplitude, while the patient with the highest baseline mean ICP (Patient 1) did not have the largest preoperative ventricular volume. In contrast, there was a possible positive trend between corrected age at surgery and baseline pulsation amplitude (Figure 6B), though the small sample size precludes any formal conclusion. Patients 3 and 5 each had exaggerated cortical subarachnoid spaces, but this did not correlate with either physiologic response or clinical outcome. Also, when estimating the likelihood of ETV/CPC success at one year, using a previously published algorithm based on corrected age and FOHR [21] there was no apparent correlation between likelihood of success and intra-operative responses. Patient 5 had the lowest predicted chance of success (about 50%) while patients 2, 3, and 4 had the highest (all >75%).

**Figure 6.**
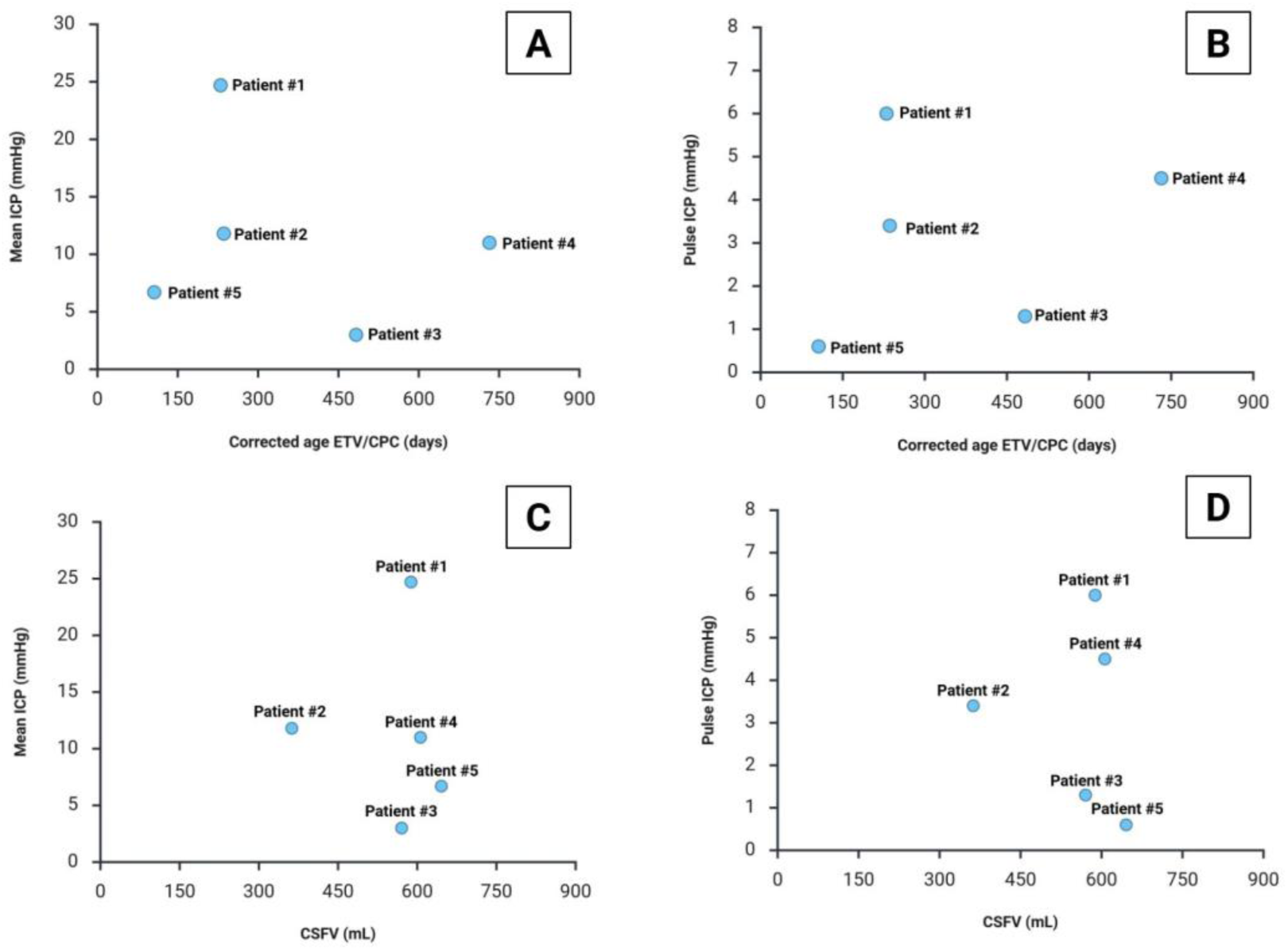
Scatter plot of (A) mean ICP at baseline and corrected age at surgery; (B) pulse ICP at baseline and corrected age at surgery; (C) Mean ICP at baseline and preoperative CSFV; and (D) Pulse ICP at baseline and preoperative CSFV. ICP: intracranial pressure; CSFV: cerebrospinal fluid volume.

## DISCUSSION

This proof-of-concept study represents the first direct, intraoperative human measurements of concurrent ventricular pressure and pulsation amplitude during ETV/CPC for infant hydrocephalus. Using a minimally invasive fiber-optic ICP monitoring method integrated passively into the operative ventriculoscope, we demonstrated that real-time measurement of mean and pulsatile ICP throughout the ETV/CPC procedure is both feasible and safe. If validated in larger cohorts, this methodology could provide a direct intraoperative window into individual physiological responses with potential implications for surgical decision-making; however, the current dataset is insufficient to support that inference, and all five patients were ultimately treated successfully by ETV/CPC.

ETV/CPC is the only established effective alternative to ventricular shunts in infant hydrocephalus; yet 30–40% of patients treated before 12 months of age will ultimately fail [4], and the physiological determinants of failure have not been established. According to the hydrodynamic model[5], ETV functions as a pulsation absorber reducing intraventricular pulsation amplitude, while CPC further attenuate pulsation amplitude by reducing the choroid plexus role as a pulsation driver [13]. This study was designed to test that two-step hypothesis through direct, real-time measurement of ICP and pulsation amplitude during ETV/CPC.

This theoretical mechanism is broadly supported by our results: Four of five patients demonstrated a substantial decrease in mean ICP (56–97%) by the end of the combined procedure. Patients 1 and 3 showed the anticipated stepwise reductions in both mean ICP and pulsation amplitude following each procedural component, most directly consistent with the hydrodynamic model. Patient 4 exhibited a large drop in mean ICP after ETV (56%) with a modest decrease in pulsation amplitude, neither of which was further augmented by CPC. Patient 5 had negligible baseline pulsation amplitude but demonstrated substantial sequential decreases in mean ICP following ETV and then CPC. The response patterns in these four patients are broadly consistent with the hydrodynamic model, however, the inter-patient variability suggests that the mechanisms driving hydrocephalus and its treatment by ETV/CPC are likely multifactorial and not fully explained by alterations in ICP alone.

Patient 2 was a notable outlier in both intraoperative physiology and clinical course. No meaningful change in mean ICP or pulsation amplitude was observed following either ETV or CPC; pulsation amplitude in fact increased after ETV and remained above baseline at procedure end. Clinically, ventriculomegaly and head circumference z-score continued to increase for the first three months postoperatively, raising concern for treatment failure despite an asymptomatic child with a sunken fontanel. Shunt placement was deferred before stabilizing at four months. Whether the simultaneous fenestration of a prepontine arachnoid cyst influenced the intraoperative ICP responses or the subsequent clinical trajectory cannot be determined from available data. The correspondence between this patient’s absent intraoperative ICP response and atypical clinical course is intriguing but circumstantial. Whether intraoperative physiological responses have predictive value for ETV/CPC outcome remains an open question requiring larger cohorts that include definitive treatment failures to determine the merit of this conjecture.

A notable and unexpected finding in this study was that baseline mean ICP was relatively low in four of five patients (Figure 5A) despite significant ventriculomegaly and a clinical picture of progressive hydrocephalus. In addition, there was no apparent correlation between baseline ICP and preoperative CSFV across patients (Figure 5C–D). Interpreting these observations requires acknowledging that intracranial compliance, the capacity of the intracranial compartment to accommodate volume changes without a proportionate rise in pressure, differs substantially in infants compared to older individuals for two distinct reasons. First, prior to cranial suture fusion, increases in intracranial volume are partially accommodated by outward expansion of the calvarium, effectively increasing total intracranial capacity rather than driving up mean pressure [29]. This manifests clinically as progressive macrocephaly, which is why head circumference and ventricular size have historically served as the primary surrogate markers of disease severity in infants rather than direct ICP measurement. Second, and independently, brain parenchyma is more viscoelastic and deformable in infants than in older children with tissue stiffness increasing progressively with age due to myelination [12]. This high parenchymal deformability means that repeated intraventricular pulsatile forces, delivered by the choroid plexus with each cardiac cycle, cause progressive displacement of surrounding brain tissue, allowing the ventricles to expand outward even at relatively low mean pressure [5, 10, 11, 30, 31]. Mathematical modeling of age-dependent tissue stiffness suggested that infant parenchyma undergoes substantially greater displacement than adult tissue under equivalent pulsatile ventricular loading [30, 31], and in vivo MR elastography corroborates this mechanism, demonstrating that brain tissue softening, most pronounced in the periventricular region, is associated with progressive ventricular expansion even at normal mean ICP [32]. These two contributors to high intracranial compliance in infancy are not interchangeable in their clinical implications: the expansile calvarium explains why mean ICP is buffered despite large ventricular volumes, while the deformable parenchyma explains why pulsatile rather than mean pressure may be the primary driver of ventricular expansion, and why younger infants, with the highest parenchymal deformability, are more vulnerable to ETV/CPC failure. However, it remains unclear how ventricular expansion and progressive compression of the surrounding brain parenchyma affect intracranial compliance during the evolution of infant hydrocephalus

These mechanisms together account for the dissociation between ICP and ventricular volume observed in this study and reported previously in the hydrocephalus literature [33–35]. Prior investigations have documented that mean ICP does not reliably predict ventricular volume in hydrocephalus [34], that direct continuous ICP monitoring reveals no proportional correlation between ventricular volume and either static or pulsatile ICP in non-communicating hydrocephalus [35], and that mean ICP is not always elevated in infants with substantial ventriculomegaly [33]. In the present study, this is most relevant to Patient 5’s response: as the youngest patient and the one with the highest preoperative CSFV z-score, Patient 5 had the lowest baseline pulsation amplitude of all patients. This is consistent with high baseline intracranial compliance at young age: the parenchyma deforms so readily under pulsatile loading that little pressure amplitude accumulates within the ventricular cavity, even as the ventricles expand progressively. The large reduction in mean ICP following CPC relative to ETV in this patient further suggests that choroid plexus reduction provided the additional dampening that ETV alone could not in this high-compliance, high-volume setting, consistent with the established observation that younger age is one of the strongest predictors of ETV/CPC failure [4, 15, 21, 22], and with the hypothesis that high intracranial compliance renders the pulsation absorber effect of ETV alone insufficient in the youngest patients [5, 12, 22]. Taken together, these findings highlight that in infant hydrocephalus, neither ventricular size nor mean ICP alone reliably captures the underlying pathological mechanism. Importantly, Patient 5 further illustrates that even pulsatile ICP may be dampened to negligible levels in the youngest and most compliant patients, where parenchymal deformability absorbs pulsatile forces before they register as measurable pressure amplitude. This underscores the need for direct compliance measurement as an informative physiological assessment in this population.

Direct measurement of intracranial compliance through intraoperative CSF infusion testing represents the logical next step in this research. Infusion testing characterizes the pressure-volume relationship of the entire CSF space, from which resistance to CSF outflow and overall system elasticity can be derived according to Marmarou’s model [36, 37]. In infants, this composite measure is substantially shaped by brain parenchymal deformability making it the closest available approximation of the parenchymal compliance we hypothesize to be a determinant of ETV/CPC outcome, while also capturing CSF outflow dynamics that may independently influence treatment response. Building on the methodology described here, future studies combining intraoperative ICP monitoring with direct compliance assessment will allow testing of whether intracranial compliance thresholds predict both the intraoperative ICP response and longer-term ETV/CPC outcome.

Finally, it should be noted that the interpretation of baseline ICP values in this study is constrained by the absence of validated normative reference values for mean and pulsatile ICP in infants. Because direct ICP measurement requires an invasive procedure, available pediatric data derive predominantly from pathological populations [38–40], making it challenging to define what constitutes a physiologically normal mean or pulsatile ICP at a given age. The characterization of baseline ICP as “low” in this study is therefore relative rather than absolute. Establishing age-specific normative ICP parameters in infancy, ideally through non-invasive proxies validated against direct measurements, represents a critical unmet need that is essential for placing intraoperative measurements of the kind described here into proper physiological context.

## CONCLUSIONS

This preliminary investigation provides the first direct intraoperative human evidence of mean and pulsatile ICP changes during ETV/CPC in infant hydrocephalus. Continuous ICP measurement throughout the procedure was feasible and safe using a fiber-optic sensor integrated passively into the operative ventriculoscope. In three out of five patients, the observed physiological responses were consistent with the proposed hydrodynamic therapeutic mechanism of ETV/CPC, while the one outlier demonstrated an atypical clinical course. Despite all patients having a clinically successful ETV/CPC in the follow-up, the inter-patient variability in the physiological responses indicates that the mechanisms underlying hydrocephalus and its treatment by ETV/CPC are likely more complex and remain incompletely understood. These findings warrant further investigation in larger prospective cohorts to clarify the factors governing intraoperative physiological responses and their relationship to clinical outcomes.

## DECLARATIONS

### Ethics approval and consent to participate

This study received ethical approval from the institutional review board at Boston Children’s Hospital. The guardians of all included patients provided written consent to participate in this study. All research procedures were conducted following the ethical principles of the Declaration of Helsinki.

### Consent for publication

Not applicable.

### Availability of data and materials

The datasets used in the current study are available from the corresponding author on reasonable request.

## Funding

NIH 5R01HD096693

### Authors’ Contributions

*Concept and design:* BCW, JS, and PL.

*Acquisition, analysis, and interpretation:* all authors.

*Drafting of the manuscript:* MHY, BCW, and PL.

*Critical review of the manuscript for important intellectual content:* all authors.

*Statistical analysis:* not applicable.

*Administrative, technical, or material support:* PEG, JS, BCW, and PL.

*Supervision:* JS, BCW, and PL.

## Data Availability

All data produced in the present study are available upon reasonable request to the authors.

## ABBREVIATIONS

ETV: endoscopic third ventriculostomy
CPC: choroid plexus cauterization
ETV/CPC: endoscopic third ventriculostomy combined with choroid plexus cauterization
CSF: cerebrospinal fluid
ICP: intracranial pressure
CSFV: cerebrospinal fluid volume
FOHR: frontal and occipital horn ratio
BPV: brain parenchymal volume

## Acknowledgements

We thank Christine Benson and Ashley Birch for their invaluable administrative and clinical support in coordinating and implementing the infrastructure necessary for the intraoperative ICP monitoring.

## REFERENCES

[1] Drake JM, Kestle JR, Milner R, Cinalli G, Boop F, Piatt J Jr, Haines S, Schiff SJ, Cochrane DD, Steinbok P, MacNeil N. Randomized trial of cerebrospinal fluid shunt valve design in pediatric hydrocephalus. Neurosurgery. 1998 Aug;43(2):294–303; discussion 303-5. doi: 10.1097/00006123-199808000-00068. PMID: 9696082.

[2] Piatt JH Jr, Cosgriff M. Monte Carlo simulation of cerebrospinal fluid shunt failure and definition of instability among shunt-treated patients with hydrocephalus. J Neurosurg. 2007 Dec;107(6 Suppl):474–8. doi: 10.3171/PED-07/12/474. PMID: 18154016.

[3] Kulkarni AV, Drake JM, Kestle JR, Mallucci CL, Sgouros S, Constantini S; Canadian Pediatric Neurosurgery Study Group. Endoscopic third ventriculostomy vs cerebrospinal fluid shunt in the treatment of hydrocephalus in children: a propensity score-adjusted analysis. Neurosurgery. 2010 Sep;67(3):588–93. doi: 10.1227/01.NEU.0000373199.79462.21. PMID: 20647973.

[4] Warf BC, Weber DS, Day EL, Riordan CP, Staffa SJ, Baird LC, Fehnel KP, Stone SSD. Endoscopic third ventriculostomy with choroid plexus cauterization: predictors of long-term success and comparison with shunt placement for primary treatment of infant hydrocephalus. J Neurosurg Pediatr. 2023 May 12;32(2):201–213

[5] Warf BC. Congenital idiopathic hydrocephalus of infancy: the results of treatment by endoscopic third ventriculostomy with or without choroid plexus cauterization and suggestions for how it works. Childs Nerv Syst. 2013 Jun;29(6):935–40. doi: 10.1007/s00381-013-2072-1. Epub 2013 Mar 13. PMID: 23483331.

[6] Warf BC. Three steps forward and 2 steps back: the Echternach procession toward optimal hydrocephalus treatment. Neurosurgery. 2014 Aug;61 Suppl 1:105–10. doi: 10.1227/NEU.0000000000000376. PMID: 25032538.

[7] Dandy WE. EXPERIMENTAL HYDROCEPHALUS. Ann Surg. 1919 Aug;70(2):129–42. doi: 10.1097/00000658-191908000-00001. PMID: 17864139; PMCID: PMC1410318.

[8] Oi S, Di Rocco C (2006) Proposal of “evolution theory in cere– brospinal fluid dynamics” and minor pathway hydrocephalus in developing immature brain. Childs Nerv Syst 22:662–669

[9] Bateman GA, Brown KM (2012) The measurement of CSF flow through the aqueduct in normal and hydrocephalic children: from where does it come, to where does it go? Childs Nerv Syst 28:55–63

[10] Di Rocco C, Pettorossi VE, Caldarelli M, Mancinelli R, Velardi F (1978) Communicating hydrocephalus induced by mechanically increased amplitude of the intraventricular cerebrospinal fluid pressure: experimental studies. Exp Neurol 59(1):40–52

[11] Pettorossi VE, Di Rocco C, Mancinelli R, Caldarelli M, Velardi F (1978) Communicating hydrocephalus induced by mechanically increased amplitude of the intraventricular cerebrospinal fluid pulse pressure: rationale and method. Exp Neurol 59(1):30–39

[12] Wilkie KP, Drapaca CS, Sivaloganathan S. Aging impact on brain biomechanics with applications to hydrocephalus. Math Med Biol. 2012 Jun;29(2):145–61. doi: 10.1093/imammb/dqr001. Epub 2011 Mar 10. PMID: 21393375.

[13] Bering EA Jr. Circulation of the cerebrospinal fluid: demonstration of the choroid plexuses as the generator of the force for flow of fluid and ventricular enlargement. J Neurosurg. 1962;19(5):405–413.

[14] Kulkarni AV, Schiff SJ, Mbabazi-Kabachelor E, Mugamba J, Ssenyonga P, Donnelly R, Levenbach J, Monga V, Peterson M, MacDonald M, Cherukuri V, Warf BC. Endoscopic Treatment versus Shunting for Infant Hydrocephalus in Uganda. N Engl J Med. 2017 Dec 21;377(25):2456–2464. doi: 10.1056/NEJMoa1707568. PMID: 29262276; PMCID: PMC5784827.

[15] Warf BC. Comparison of endoscopic third ventriculostomy alone and combined with choroid plexus cauterization in infants younger than 1 year of age: a prospective study in 550 African children. J Neurosurg [Internet]. 2005 Dec;103(6 Suppl):475–81. Available from: http://thejns.org/doi/abs/10.3171/ped.2005.103.6.0475

[16] Warf BC, Tracy S, Mugamba J. Long term outcome for combined endoscopic third ventriculostomy and choroid plexus cauterization (ETV-CPC) compared to ETV alone for congenital aqueductal stenosis in African infants. J Neurosurg Pediatr. 2012;10(2):108–111.

[17] Warf BC, Campbell JW. Combined endoscopic third ventriculostomy and choroid plexus cauterization (ETV/CPC) as primary treatment of hydrocephalus for infants with myelomeningocele: long-term results of a prospective intention to treat study in 115 African infants. J Neurosurg Pediatr. 2008;2(5):310–316.

[18] Warf BC, Stagno V, Mugamba J. Encephalocele in Uganda: Ethnic variation in lesion location, endoscopic management of hydrocephalus, and survival in 110 consecutive children. J Neurosurg Pediatr. 2011;7(1):88–93.

[19] Warf BC, Campbell JW, Riddle E. Initial experience with combined endoscopic third ventriculostomy and choroid plexus cauterization for post-hemorrhagic hydrocephalus of prematurity: the importance of pre-pontine cistern status and the predictive value of FIESTA MRI imaging. Childs Nerv Syst. 2011;27(7):1063–1071.

[20] Warf BC, Dewan M, Mugamba J. Management of Dandy-Walker Complex-associated infant hydrocephalus by combined endoscopic third ventriculostomy and choroid plexus cauterization (ETV/CPC). J Neurosurg Pediatr. 2011;8(4):377–383.

[21] Hartman E, Puglisi E, See AP, Stone SSD, Warf BC. Predicting endoscopic third ventriculostomy with choroid plexus cauterization success with machine learning: the importance of ventricle size and the irrelevance of etiology. J Neurosurg Pediatr. 2026 May 8:1–7. doi: 10.3171/2026.1.PEDS25684. Epub ahead of print. PMID: 42102402.

[22] Yoshikawa MH, Yu M, Tatz J, Tran H, Feldman HA, Luviano AS, Grant PE, Sutin J, Warf BC, Lin PY. Cerebrospinal fluid volume outperforms traditional measures for predicting treatment failure in infant hydrocephalus. Fluids Barriers CNS. 2026 May 23. doi: 10.1186/s12987-026-00819-0. Epub ahead of print. PMID: 42177495.

[23] Wilson CB, Bertan V. Interruption of the anterior choroidal artery in experimental hydrocephalus. Arch Neurol. 1967;17(6):614–619.

[24] Eide PK. The correlation between pulsatile intracranial pressure and indices of intracranial pressure-volume reserve capacity: results from ventricular infusion testing. J Neurosurg. 2016 Dec;125(6):1493–503.

[25] Yu M, Yoshikawa MH, Sosa Luviano A, Schiff SJ, Monga V, Warf BC, et al. Automated brain and CSF volume assessment in infant hydrocephalus using deep learning [preprint]. medRxiv. 2026 May 7:2026.05.07.26352592. doi:10.64898/2026.05.07.26352592.

[26] Paul A. Yushkevich, Joseph Piven, Heather Cody Hazlett, Rachel Gimpel Smith, Sean Ho, James C. Gee, and Guido Gerig. User-guided 3D active contour segmentation of anatomical structures: Significantly improved efficiency and reliability. Neuroimage. 2006 Jul 1; 31(3):1116–28.

[27] Peterson MR, Cherukuri V, Paulson JN, et al. Normal childhood brain growth and a universal sex and anthropomorphic relationship to cerebrospinal fluid. J Neurosurg Pediatr. 2021 Jul 9;28(4):458–468. doi: 10.3171/2021.2.PEDS201006.

[28] Rigby RA, Stasinopoulos DM. Smooth centile curves for skew and kurtotic data modelled using the Box-Cox power exponential distribution. Stat Med. 2004 Oct 15;23(19):3053–76. doi: 10.1002/sim.1861. PMID: 15351960.

[29] Hladky SB, Barrand MA. Regulation of brain fluid volumes and pressures: basic principles, intracranial hypertension, ventriculomegaly and hydrocephalus. Fluids Barriers CNS. 2024 Jul 17;21(1):57. doi: 10.1186/s12987-024-00532-w. PMID: 39020364; PMCID: PMC11253534.

[30] Sivaloganathan S, Stastna M, Tenti G, Drake J. A viscoelastic model of the brain parenchyma with pulsatile ventricular pressure. Appl Math Comput. 2005;165:687–698

[31] Wilkie KP, Drapaca CS, Sivaloganathan S. A mathematical investigation of the role of intracranial pressure pulsations and small gradients in the pathogenesis of hydrocephalus. Appl Math Comput. 2012;218(18):9505–9519

[32] Streitberger KJ, Wiener E, Hoffmann J, Freimann FB, Klatt D, Braun J, Lin K, McLaughlin J, Sprung C, Klingebiel R, Sack I. In vivo viscoelastic properties of the brain in normal pressure hydrocephalus. NMR Biomed. 2011 May;24(4):385–92. doi: 10.1002/nbm.1602. Epub 2010 Oct 7. PMID: 20931563.

[33] Flanders TM, Lang SS, Ko TS, Andersen KN, Jahnavi J, Flibotte JJ, Licht DJ, Tasian GE, Sotardi ST, Yodh AG, Lynch JM, Kennedy BC, Storm PB, White BR, Heuer GG, Baker WB. Optical Detection of Intracranial Pressure and Perfusion Changes in Neonates With Hydrocephalus. J Pediatr. 2021 Sep;236:54–61.e1. doi: 10.1016/j.jpeds.2021.05.024. Epub 2021 May 15. PMID: 34004191; PMCID: PMC8403158.

[34] Børgesen SE, Gjerris F. Relationships between intracranial pressure, ventricular size, and resistance to CSF outflow. J Neurosurg. 1987 Oct;67(4):535–9. doi: 10.3171/jns.1987.67.4.0535. PMID: 3655891.

[35] Eide PK, Sorteberg A, Sorteberg W. Association between ventricular volume measures and pulsatile and static intracranial pressure scores in non-communicating hydrocephalus. Acta Neurochir. 2015;157(3):369–78. PMID: 25680584

[36] Czosnyka M, Wollk-Laniewski P, Batorski L, Zaworski W. Analysis of intracranial pressure waveform during infusion test. Acta Neurochir. 1988;93(3-4):140–5.

[37] Katzman R, Hussey F. A simple constant-infusion manometric test for measurement of CSF absorption. I. Rationale and method. Neurology. 1970 Jun;20(6):534–44.

[38] Pedersen SH, Lilja-Cyron A, Andresen M, Juhler M. The Relationship Between Intracranial Pressure and Age-Chasing Age-Related Reference Values. World Neurosurg. 2018 Feb;110:e119–e123. doi: 10.1016/j.wneu.2017.10.086. Epub 2017 Oct 26. PMID: 29107158.

[39] Pedersen SH, Lilja-Cyron A, Astrand R, Juhler M. Monitoring and Measurement of Intracranial Pressure in Pediatric Head Trauma. Front Neurol. 2020 Jan 14;10:1376. doi: 10.3389/fneur.2019.01376. PMID: 32010042; PMCID: PMC6973131.

[40] Sæhle T, Eide PK. Intracranial pressure monitoring in pediatric and adult patients with hydrocephalus and tentative shunt failure: a single-center experience over 10 years in 146 patients. J Neurosurg. 2015 May;122(5):1076–86. doi: 10.3171/2014.12.JNS141029. Epub 2015 Feb 13. PMID: 25679270.

